# Understandings of ultra-processed foods: a qualitative interview study of UK adults with responsibility for household food activities

**DOI:** 10.1101/2025.08.18.25332943

**Authors:** Michael Essman, Jennie C Parnham, Kiara Chang, Mike Etkind, Prisha Shah, Deb Smith, Milica Vasiljevic, Emma Boyland, Steven Cummins, Eszter P Vamos, Martin White, Jean Adams

## Abstract

**Background:** There is accumulating evidence that ultra-processed foods are associated with many non-communicable diseases. In the United Kingdom, ultra-processed foods account for more than half of mean daily energy intake. There is limited evidence describing how individuals make sense of ultra-processed foods in their daily lives. We aimed to explore: public perceptions of ultra-processed foods and their relationship to health; how these are shaped by different information sources; whether perceptions influence purchasing and consumption; and proposed solutions for reducing ultra-processed food consumption.

**Methods:** We undertook 30 qualitative, one-to-one interviews with people living in the UK, aged 18 years or older, with household responsibility for food activities recruited using social media advertisements. Interviews followed a semi-structured topic guide reflecting the aims. Data were analysed using reflexive thematic analysis. Members of a Public Involvement and Engagement group helped interpret emergent findings.

**Results:** Food processing often made more sense to participants as a continuum rather than ordinal categories. There was particular confusion regarding the boundary between processed and ultra-processed food.

Participants described an overwhelming food-related information environment. Personal and anecdotal experience was particularly powerful. While industry-sponsored information was met with scepticism, the personal testimonies of social media influencers were seen as relatable or helpful.

Participants’ perceptions of ultra-processed foods sometimes influenced their purchasing and consumption. However, cost, convenience, taste and family practices were often as or more important.

Approaches to help reduce UPF consumption ranged from education to fiscal policies.

**Conclusions:** These findings suggest that the public need help navigating the complex food information landscape and not just with filling specific knowledge gaps around ultra-processed foods and other food-related topics. Policy approaches should combine improved public communication with upstream interventions that modify the food environment to make choosing less processed foods easier and more appealing.

## BACKGROUND

The increasing consumption of harmful commodities, including tobacco, alcohol, and unhealthy foods, has been recognised as a major driver of the global burden of non- communicable diseases (1). Among unhealthy foods, there is accumulating evidence that ultra-processed foods (UPFs) are associated with poor diet quality, increased risk of many non-communicable diseases, and greater mortality (2–7).

The term UPF was introduced in 2009 as part of the Nova classification, which categorises foods by the extent and purpose of industrial processing (8,9): group 1 are unprocessed or minimally processed foods, group 2 are culinary ingredients, group 3 are processed foods and group 4 are UPFs. UPFs are industrially formulated products designed for convenience, hyper-palatability, and commercial profitability (8). In the United Kingdom (UK), UPFs account for more than half of the daily energy intake of adults, with even higher intakes among children and adolescents (10–13). This widespread consumption is entrenched by recent food price inflation and marketing practices that promote UPFs as affordable and convenient options (14,15). Overall, 41% of European consumers report that UPFs are more convenient than minimally processed foods (16).

Despite their widespread consumption, public understanding of what constitutes a UPF remains limited and perceptions mixed. For example, a study of 2,386 UK adults found that while 73% were aware of the term ‘UPF’, participants on average correctly categorised 54% of UPF items and only 13% of participants accurately categorised all UPFs (17). Another UK report found that although perceptions of UPFs are generally negative, there are varying levels of understanding and concern, with some participants considering them unavoidable (18). Similarly, although 65% of Europeans believe that UPFs are unhealthy, only 56% reported trying to avoid them (16).

This limited public understanding is likely shaped by a complex and often contradictory information environment. In the contemporary digital environment – characterised by user- generated content and social media – health information is increasingly shared through peer networks (19), which can be difficult to navigate and include misinformation (20). Research on nutrition misinformation suggests that food choices and dietary beliefs are especially vulnerable to conflicting messages because social media algorithms amplify both scientific evidence and industry narratives (21). This creates a polarised information landscape in which individuals are frequently exposed to contradictory and false claims, making it difficult to separate credible guidance from industry marketing, scientifically unproven positions or personal anecdotes (22,23). There is also a lack of transparency regarding the conflicts of interest and credentials of online sources (24). In particular, the food industry employs a set of tactics, both on and offline, to manipulate consumer demand including deceptive health and nutrition claims to influence food and health policy debates (25,26). The food industry also uses celebrities and online ‘influencers’ to promote unhealthy products, and studies have demonstrated that children and adolescents are particularly susceptible to these marketing tactics (27–29).

This complex information space interacts with debates occurring in the scientific community including whether the Nova classification system has adequate specificity and inter-rater reliability – although many studies have reported good inter-rater reliability – and whether associations with health outcomes are causal (13,30,31). In the context of rising food insecurity in the UK, there are also equity concerns about demonising a broad and heterogeneous category of affordable and accessible foods (32).

Despite the emerging scientific evidence that UPFs are generally deleterious to health, uncertainty regarding whether the public understands the concept of UPF remains unanswered (33). Notably, a recent study from Brazil, where the UPF concept originated, reported that most Brazilians found the term confusing (34). Understanding public perceptions of UPFs is important because policies are more likely to have public support and behavioural impact if the public understands the rationale behind them and perceives them as beneficial (35). Currently, there is limited evidence describing how individuals make sense of UPFs in their daily lives. The aims of this study were first, to explore public understanding of the concept of UPFs and perceptions of the relationship between UPFs and health; second, to explore how these perceptions were shaped by different information sources; third, to explore whether perceptions of UPFs influence purchasing and consumption; and fourth, to explore proposed solutions for reducing UPF consumption. We focused on individuals with responsibility for household food activities assuming they were more likely to be making everyday decisions about UPFs.

## METHODS

This study is reported as per the Consolidated Criteria for Reporting Qualitative Research guidelines (Supplementary File 1 contains a completed checklist) (36).

### Study Design and Participants

We undertook qualitative, one-to-one interviews with people living in the UK, aged 18 years or older, who responded yes to: “Are you usually the main person in your household in charge of activities related to food, for example buying and cooking?” Participants were recruited using social media advertisements and were not previously known to the research team. A participant information sheet was provided explaining that the study aimed to understand experiences and understanding of UPFs and that interviews would be conducted by a professional researcher (Supplementary File 2). Sampling was open and iterative: while there were no fixed quotas, we monitored recruitment to support diversity across key demographic characteristics of gender, age, ethnicity, geographic location, and education.

As interviews progressed, we identified a need to increase sampling of men and made targeted adjustments to recruitment advertisements to improve gender balance. Recruitment was halted after 30 interviews, with consistent themes emerging across participants.

### Data Collection

Interviews were conducted in July-October 2024 via video conference. ME conducted interviews with participants and each participant took part in one interview. ME is a male researcher who has previously attended formal qualitative research training, gained informal insight into interview technique via shadowing experienced researchers, and conducted previous qualitative interview studies. No-one else was invited to attend interviews, but other people may have been off-screen in participant locations. Each interview lasted up to 60 minutes and followed a semi-structured topic guide designed to explore participants’ understanding of UPFs (as described by the Nova classification), their relationship to health, and relevant information sources (Supplementary File 3). The topic guide was not formally piloted but was iteratively developed between participants. To elicit participants’ own conceptualisations of UPFs, interviews included a structured activity in which participants were shown a set of images depicting foods across the Nova classification. Participants were first asked to categorise the foods based on their own understanding of what constitutes a UPF. Only after this did the interviewer clarify any misconceptions, allowing for reflection on differences between participants’ prior assumptions and the formal Nova classification. All interviews were digitally audio-recorded with participants’ consent, transcribed verbatim, and anonymised. Transcripts were not returned to participants for checking and they were not invited to feedback on the findings. As a gesture of appreciation for their time, participants received a £20 online shopping voucher.

### Data Analysis

Data were analysed using reflexive thematic analysis (37–39). This included familiarisation with the data through repeated reading, coding, searching for themes, reviewing and refining themes, and producing a final analysis. Reflexive thematic analysis was chosen for its flexibility in identifying patterns while allowing for detailed exploration of participants’ views. While existing literature on UPFs informed our interview topic guide, our analytical approach was inductive.

NVivo version 12 was used for data organisation, and themes were iteratively refined using whiteboard diagramming to examine relationships between them. The coding process included both explicit meanings of participant statements as well as researcher interpretation to explore underlying assumptions, social influences, and structural factors shaping participants’ perspectives. Anonymised verbatim quotations were selected to illustrate key findings.

To enhance the rigour and trustworthiness of the analysis, three members of the research team (ME, JP, JA) independently read and coded a subset of transcripts, then met in a data clinic to compare interpretations and discuss emerging themes and assumptions.

Additionally, several rounds of sense-making were conducted with a Public Involvement and Engagement (PIE) group who also contributed to thematic development and interpretation of results, helping refine key themes and highlight gaps in understanding. The PIE group reviewed emerging themes for coherence with participant narratives, assessed the accuracy of theme definitions, and highlighted surprising or misaligned quotes.

### Reflexivity statement

This work was conducted by an inter-disciplinary team of university-based researchers in a range of research and academic positions with higher degrees (PhDs and MDs) and PIE contributors. We have expertise in dietary public health and epidemiology, evaluation of dietary public health interventions, analysis of the relationship between UPF and health outcomes and qualitative methods. PIE members have experience of contributing to a range of research on food and other topics. Our personal and professional experiences have informed our understanding of the broader food environment and the structural factors that shape dietary behaviours. Within the research team, we have a range of views on the strength of evidence on the links between UPF and health outcomes and the necessity for UPF-specific policy.

## RESULTS

Thirty people took part (Table 1). As we do not know how many people saw the social media recruitment advertisements, we do not know how many chose not to take part or why. Most participants were women (n=22, 73%), reflecting ongoing gendering of food responsibility in the UK (40). The sample was disproportionately of higher educational status (n=19, 63%) compared to about one-third in the population as a whole (41). However, the sample was diverse in terms of ethnicity (n=14, 47% White British) and region of residence. UK regions included West Midlands, East of England, South East, Wales, South West, North West, London, Scotland and Yorkshire and the Humber. One participant did not share their demographic data.

**Table 1.** Demographic characteristics of interview participants (n=30)

| ID | Age (years) | Gender | Ethnicity | Education* |
| --- | --- | --- | --- | --- |
| 1 | 70+ | Female | White British | Low |
| 2 | Not recorded | Male | Not recorded | Not recorded |
| 3 | 60-69 | Female | White non-British | High |
| 4 | 20-29 | Male | White non-British | High |
| 5 | 30-39 | Female | White non-British | High |
| 6 | 30-39 | Female | White British | High |
| 7 | 40-49 | Male | White non-British | High |
| 8 | 60-69 | Female | White British | High |
| 9 | 40-49 | Female | Asian | Medium |
| 10 | 50-59 | Female | White British | Medium |
| 11 | 60-69 | Female | White British | High |
| 12 | 40-49 | Female | Asian | Low |
| 13 | 20-29 | Female | Asian | High |
| 14 | 40-49 | Female | Asian / White British | High |
| 15 | 60-69 | Female | White British | High |
| 16 | 40-49 | Female | Asian | Low |
| 17 | 50-59 | Female | White British | Medium |
| 18 | 60-69 | Male | White British | High |
| 19 | 50-59 | Female | White British | High |
| 20 | 40-49 | Male | White British | Low |
| 21 | 40-49 | Female | White British | Medium |
| 22 | 20-29 | Female | Asian | High |
| 23 | 20-29 | Female | White non-British | High |
| 24 | 30-39 | Female | White British | High |
| 25 | 30-39 | Male | White British | High |
| 26 | 20-29 | Male | White British | Low |
| 27 | 40-49 | Male | Asian | High |
| 28 | 20-29 | Female | Asian | High |
| 29 | 20-29 | Female | Asian | Low |
| 30 | 20-29 | Female | Asian | High |

\*Education level: high = university degree and above; low = school level only; medium = anything in between.

Thematic analysis identified five interconnected themes: (1) Understanding of the concept of UPF, (2) Influences on understanding of the concept of UPF, (3) Decision making around UPFs, (4) Barriers & enablers to reduced UPF consumption, and (5) Proposed solutions to help reduce UPF consumption. Below, each theme is first described in brief followed by a more detailed discussion of sub-themes supported by illustrative quotations. Table 2 summarises the themes.

**Table 2.** Study themes, including descriptions and key quotes.

| Themes | Description | Key illustrative quotes |
| --- | --- | --- |
| Theme 1:<br>Understanding of the concept of UPFs | Participants varied in their understanding of UPFs and their ability to identify them. Many participants relied on packaging cues, branding, or ingredient lists to determine whether a food was UPF. Simplified frameworks (e.g. "single ingredient foods") were sometimes preferred to boundaries like level of processing. | I think most people understand it by nature, but I do think it maybe doesn't come to the top of mind for people as much as other terms. It's quite long, maybe it's slightly got more scientific feel. I think what I'm hearing more now is this single ingredient foods. (Participant 5)<br>If I look at a product in the supermarket and it has a list of ingredients that you generally wouldn't find in your kitchen, then they tend to be ultra-processed. So things like emulsifiers, things that make things have nice smooth consistencies. It's a chemical... been chemically processed rather than being naturally existing (Participant 11) |
| Theme 2:<br>Influences on understanding of the concept of UPF | Media, marketing, and social norms shaped participants' perceptions of UPFs, reinforcing their acceptability and desirability. While some participants were sceptical of industry messaging, others found it difficult to filter reliable information from the overwhelming volume of food-related content in advertising and social media. | I see all the advertisements, even on YouTube now they have advertisements all the time. (Participant 3)<br>For me it's seeing what my children give their children now and how it's changed so much from like my kids... I mean it's shocking the sugar that's in some of the children's foods, the baby foods, that are marketed for children. (Participant 7) |
| Theme 3:<br>Decision making around UPFs | Participants described a range of behaviours related to their beliefs about UPFs, from reducing consumption to accepting UPFs as unavoidable. Many engaged in self-experimentation, testing how reducing or eliminating certain UPFs affected their well-being. Health concerns, cultural traditions and distrust of industry marketing all motivated some participants to limit or avoid UPFs. | I think that does show that [UPFs] can be bad compared to less processed foods. And I think it's not just eczema, I think other people have said that when they eat more ultra-processed foods they become more tired... although there's not really good scientific evidence I think because there's so many stories, it does have some kind of proof to it. (Participant 13) |
| Theme 4:<br>Barriers & enablers to reduced UPF consumption | Affordability, availability, convenience, and time were major barriers to reducing UPF consumption. Some participants highlighted that fresh, minimally processed foods were becoming more expensive, while UPFs remained cheap, widely accessible, and often the more convenient option. However, some participants said discount grocery stores allowed them to avoid UPFs and save money. Food culture and upbringing influenced the ability to navigate these barriers. | Lots of people nowadays are cash-strapped and if something's cheaper and it looks good, and probably tastes quite good too, you're bringing up a family, you're going to go for the cheaper option, why wouldn't you? (Participant 15)<br>I think convenience is a big one, if you're going out somewhere to grab a chocolate bar, put it in your bag, it's pre-wrapped. Probably by the way it's been modified often it perhaps holds together a little bit more and that can be good for transportation... if you just need some sugar maybe you're not feeling great or you're a bit stressed then can help in the moment. (Participant 5) |
| Theme 5:<br>Proposed solutions for reducing UPF consumption | Participants expressed mixed views on potential policy interventions, ranging from supportive to sceptical. While many saw value in education, clear labelling, and restrictions on marketing, others emphasised the need for systemic changes, such as making healthier foods more affordable and accessible rather than simply restricting UPFs. | I think the government has got to look at the state of the nation's health, they're going big on obesity, is obesity caused by these ultra-processed meals that they're not getting enough nutrition, which is then making them eat more crap... and it's not done anything for them, it's just going to put a bigger strain on the NHS and worldwide, we're all going to suffer. (Participant 10)<br>When you enforce decisions on people they end up resenting you... to hear that the price has gone up it's like oh, for goodness sake, like just let me have my chocolate bar... I kind of get why it's being done but also it makes me feel uneasy... where does it stop? (Participant 24) |

### Theme 1: Understanding of the concept of UPFs

Participants exhibited varying levels of familiarity with, and understanding of, the term UPFs. Responses ranged from clear ‘textbook’ definitions based on the Nova classification that participants either knew or repeated, to many struggling to apply the concept consistently. Most participants reported having heard the term UPF, although six had not heard of it prior to study recruitment. While some participants were aware that UPFs were industrially processed and tend to contain multiple ingredients, many struggled to differentiate between foods in Nova group 3 (processed) and Nova group 4 (ultra-processed). Several participants expressed uncertainty about what degree of processing constituted "ultra-processing". Some participants wanted to think about a scale of processing, even within the UPF category. Participants often relied on simplified shortcuts to infer UPF status, such as ingredients they did not recognise and whether products were packaged. Some participants viewed the UPF category as overly broad and questioned whether technical classifications were useful for everyday shoppers.

#### UPFs are artificial or not natural

Some participants described UPFs as highly processed and containing additives but lacked a precise understanding of what distinguished UPFs from processed foods. Many participants identified UPFs as “artificial”, not in their "natural" state or having undergone industrial transformation. Food that was in its natural state was considered healthier.

> *You just know that if something, it’s like it’s artificial isn’t it? If something’s natural it’s good for you. If something’s been pumped with something or has been added things they’re all bad for you. It’s like anything natural. (Participant 12)*

Other alterations to food away from its ‘natural state’ were also considered by some participants to make a product UPF. For example, two participants suggested genetically modified food might be UPF.

> *Ultra processed to me means genetically modified food, it means the whole, like everything, it is not just the actual food itself. But GM food to me is also ultra processed because of the way it has been made and manufactured.* (Participant 25)

This confusion led to uncertainty about where the boundaries between minimally processed, processed, and UPFs should be drawn, with some participants struggling to determine which foods were healthier and which should be avoided. Some participants eating more restrictive diets, e.g. vegan, expressed confusion with the size of the UPF category. One recognised that many protein substitutes were classified as UPFs.

#### Reliance on packaging, ingredients, and shortcuts to identify UPFs

Due to a lack of familiarity with formal definitions, many participants used product packaging, ingredient lists, and simple shortcuts to determine whether a food was UPF. For example: the longer the ingredient list, the more likely the food was UPF.

> *I think once you start looking at the ingredients on the packet, you can quickly get a rough gauge—the more ingredients there are, the more ultra-processed it probably is.* (Participant 7)

Others followed guidelines picked up from informal sources, such as avoiding foods with many ingredients or those containing ingredients they could not recognise.

> *Something that contains ingredients that you would, you know, you just wouldn’t find in a kitchen, home,* (Participant 7)

#### Challenges in differentiating processed and ultra-processed

Distinguishing processed foods from UPFs was a particular area of confusion. Some participants described how their previous understanding of “processed food” was being reshaped by learning about UPF.

> *Obviously, I’ve heard of processed foods, but it’s the first time I’ve come across the “ultra” adjective… I’m not sure that I understand how different it is from just processed foods, I’m not sure why you would need a subdivision there, particularly. Unless there are just specific foods, like hamburgers, ice cream, etc, that fit into that sub-class.* (Participant 15)

There was uncertainty around foods perceived to be ‘borderline’, such as bread, yogurt, and plant-based alternatives, with participants expressing difficulty in determining whether a food’s level of processing was “too much.”

> *The ones that definitely obviously I’d say are the ready-meals, ultra-processed, the breakfast cereal, the cakes, the crisps. And then I would say the ones that are kind of a bit borderline for me would maybe be the supermarket bread because I know there’s quite a lot of additives in the sort of loaves you get off the shelf but again it’s bread so I wouldn’t say it was ultra-processed but I would say it’s sort of borderline perhaps.* (Participant 26)

Few participants mentioned the Nova classification system, and those who were aware of it found it somewhat arbitrary or overly complex.

### Theme 2: Influences on understanding of the concept of UPF

Perceptions of UPFs were shaped by a diverse range of influences, including traditional media, social media, advertising, and personal networks. While some participants actively sought out information on UPFs from traditional media and traditional expert sources, others engaged with UPF-related content through social media and online ‘influencers’. Participants expressed scepticism about certain sources of information, particularly those linked to the food industry, and described how conflicting messages made it difficult to determine the reliability of diet-related and UPF-specific advice. Some participants described conflicting messages between traditional experts and other sources such as social media and online ‘influencers’, making it difficult for them to determine which information was most applicable to their needs. Some participants who were most sceptical of advice from traditional experts questioned whether these individuals were also influenced by corporate interests. Marketing and advertising were felt to reinforce UPF consumption as normal and unavoidable.

#### Traditional and social media as primary sources of UPF information

For many participants, awareness of UPFs was driven by traditional media coverage, such as news articles, television documentaries, and radio discussions. After awareness was raised through traditional sources, social media then played a larger role in shaping participants’ perceptions of UPFs.

> *We’ve gone through a lot of changes with the ultra-processed food because of things that I’ve been reading online, that I’ve seen on TikTok, and I’ve actually been drawn to [actress] and I’ve read her book, so over the past year my thoughts of buying food have completely changed and I try to cook more, I avoid the low fat stuff, and get what I’d call, now call ‘proper food’, real food.* (Participant 21)

However, participants acknowledged concerns about misinformation and conflicting messages, with some describing how social media algorithms exposed them to extreme or exaggerated claims.

> *It’s full of false news, false information, I’m aware of this, and therefore I never trust one source of information, it doesn’t matter what it is about… I always try to compare at least two, three different websites for sources of information because I’ve noticed you can’t trust anyone these days without checking the source.* (Participant 6)

Other sources of information included friends or other personal relationships, as these provided relatable real-life stories.

> *My hairdresser has just gone on a non-ultra-processed food way of life, he’s lost shedloads of weight, he’s got loads of energy, and seems to be really well.* (Participant 10)

#### Scepticism toward the UPF industry and conflicting narratives

Participants expressed deep scepticism about information provided by the food industry, particularly large multinational corporations. Some viewed industry messaging as deliberately misleading, designed to downplay concerns about UPFs while promoting their products.

> *Certainly the industry’s interests are not informing you of the manufacturing processes and the ingredients they use. … if you get information from Nestlé or from other big food manufacturers or multinationals, it’s not in their interests to tell you how things are.* (Participant 2)

Others felt that corporate marketing exploited consumer confusion, making it difficult to determine whether certain products were genuinely healthier or just repackaged as “better” options.

> *I think that sometimes what annoys me is when they try to make out that it’s quite healthy but actually they try to play on our preconceptions maybe brown bread being healthy but actually there’s a lot of sugars and stuff in the bread that you could just say it’s not really healthy, it’s still ultra-processed.* (Participant 5)

This scepticism extended to traditional experts, with some participants questioning whether official recommendations were outdated or influenced by industry interests.

> *You have to be a little bit sceptical because even a doctor, you never know who he’s working with. You pay attention of course to who is saying that, but you can’t really be sure who is connected with who because it’s a very complicated world at the moment.* (Participant 4)

#### Strategic marketing and the normalisation of UPF consumption

Participants highlighted the pervasive role of marketing in shaping dietary norms, particularly through targeted promotions, supermarket placement, and fast-food delivery app notifications. UPFs were often positioned as the default or easy choice, in contrast to the implicitly difficult choice of home cooking. This marketing not only normalised UPFs but also contributed to participants’ scepticism of the food industry.

> *They are bombarding us with so many advertisements about how it’s normal to go out and eat out and order food. Because like Uber Eats for example or other applications are sending you like notifications every day, “can you order that? Why don’t you order that?” So it’s like a combination of things that’s making you think this is very normal to order from, having a delivery and relax instead of cooking.* (Participant 4)

> *To encourage people to have like a cake or a sweet, often they are in the discounted aisle or they put them near the checkouts with special deals because they know people are going to sort of crack just before they make their purchase.* (Participant 5)

Participants also discussed how marketing presented aspirational lifestyles with sports figure and celebrity endorsements influencing young people’s purchasing decisions.

> *I know from my sons they are always looking at what the footballers are wearing, because they’re both into football as well that they do notice those sort of things, whether they’re promoting a drink or a fizzy drink or crisps.* (Participant 14)

### Theme 3: Decision making around UPFs

Participants described a range of decisions in response to information about UPFs, from dietary changes and self-experimentation to continued consumption. These were highly individualised, shaped by health concerns, media exposure, social influences, and personal circumstances. Personal health concerns were an important motivator to change, whereas convenience, cost, and ingrained habits were acknowledged barriers to change.

#### Relationship between UPFs and health

Most participants described UPFs as bad for health, regardless of how much they consumed. Some expressed uncertainty around whether there was scientific consensus and the long-term consequences of UPF consumption.

> *I think the general consensus is that they are negative, end of story. I do think there’s just a bit of ambiguity surrounding what are the actual negative consequences of them though. …. We don’t know perhaps the long-term effects.* (Participant 26)

Participants expressed concern about the lack of clear guidance on UPFs, particularly regarding safe consumption levels and potential health risks, and suggested that this lack of information was a barrier to changing UPF consumption.

> *I do try and maintain the balance. I mean there’s always, my plate is always loaded with grains and nuts … but I’ve never came across any guidance. Firstly, on dangers [of] ultra-processed foods, secondly on safe, quantities: which are safer for you, or ways to mitigate any risks…. So, it just, it does just feel like I’m blind on it.* (Participant 20)

Many participants linked UPFs to non-communicable diseases. While some participants actively avoided UPFs, others were less convinced of their harmfulness, suggesting that moderation or balancing UPFs with healthier foods was the most reasonable strategy.

#### Efforts to reduce UPF consumption

Many participants reported making conscious efforts to reduce UPF consumption, often after encountering information about their potential health risks. Some shifted towards more home cooking and avoiding highly processed packaged foods, while others described incremental adjustments, such as cutting out specific types of UPFs or selectively scrutinising ingredient lists.

> *I am trying as much as possible to go for the non-processed foods… I’d definitely say avoiding them more based on wanting to improve my health and what I’ve heard on social media or on TV or in articles… and I experimented a bit with different things and I found that has helped me.* (Participant 5)

#### Self-experimentation

Several participants described experimenting with their diets based on information encountered through media or personal networks. This often took the form of informally testing hypotheses about how UPFs might affect their health.

> *I now know what to look for, for sorbitol and mannitol which are natural in some foods, but when they’re put in, in excess, that I really can’t tolerate them. You mentioned sweets earlier, I don’t touch any of those sugar free sweets, or sugar free gums, anything like that, I know that it’s really going to give me pain and I’m not going to feel well.* (Participant 10)

For many, personal health concerns played a central role in dietary changes, prompting them to experiment with elimination diets or a shift toward whole foods.

> *I’ve had a lot of issues, just felt very uncomfortable when I eat some foods. I did the [Personalised Nutrition Diet Programme] last September, which gave me some insights into what seemed to suit my body a little bit better, so I’ve kind of adopted a little bit more of that way of eating, so more fermented foods, a lot more nuts, seeds, beans.* (Participant 11)

For some participants, these trial-and-error dietary adjustments led to a deepening sense of distrust toward mainstream dietary guidance, particularly when professional recommendations contradicted their lived experience.

> *I went to a gastroenterologist and dietician and they suggested that I stopped milk which didn’t make me feel any better and since I’ve reintroduced milk and I’m fine so that’s got me thinking that some of the health advice that I’ve been given… might be a little bit dated… that’s why I kind of took it under my own steam to do more research and ultra-processed foods just kept popping up.* (Participant 21)

### Theme 4: Barriers & enablers of reduced UPF consumption

Participants described a range of structural, environmental, and cultural factors that shaped their ability to reduce UPF consumption. These included time scarcity and the perceived convenience of UPFs, price constraints, and the widespread availability of ready-made foods. Cultural background, family upbringing, and generational norms also influenced what participants considered a “normal” or acceptable diet.

#### Accessibility and convenience

Participants frequently cited convenience as a primary driver of UPF consumption, often describing UPFs in terms of saving time, avoiding stress, and fitting food preparation around other responsibilities. Some participants emphasised that time constraints often outweighed cost considerations.

> *I’d say convenience because yes, we are time poor and it is time to cook it fresh and having to do all of that, I think is a bigger factor, that’s the most important.* (Participant 9)

Many participants emphasised the ease of purchasing, preparing, and storing UPFs, which were often considered the default option for busy households.

> *If I finish work at 5 and then I have to cook something, it’s not that easy if I need to go and buy some groceries as well. So, on days that both me and my partner work and we haven’t prepared something from the previous day, it might be a bit difficult to avoid an ultra-processed food.* (Participant 3)

Some participants described how neighbourhood food availability shaped their choices making UPF consumption almost unavoidable.

> *The only benefits are that when you have no time or you’re lazy after a long day at work, this is the only food sometimes available to you, whether you buy it prepared in store or you order takeaway… around by me there are just so, so many fast-food places and you look out the window and every few seconds there’ll be a delivery driver going past… we’ve made it too easy to get hold of fast food.* (Participant 6)

#### Price

Price was also a key influence on UPF consumption often discussed alongside convenience. Some participants viewed UPFs as more affordable, while others noted that rising prices or strategic shopping could make fresh food comparable in cost. Discounted ready meals and snacks were commonly seen as a low-cost choice.

> *It’s going to be cheaper, if you go to somewhere like [discounter freezer store] and say they sometimes have those three meals for a fiver, they’re ready meals… if you were a single person and say you needed six meals right, and you picked up those you could feed yourself in £10* (Participant 9)

Others suggested that either shopping at local farmers markets, discount supermarkets, or cooking at home could offer similar prices to UPFs.

> *I tend to use the large German supermarkets because we buy a lot of food… they’ve probably increased those packaged meals, the price of them, I don’t think they’re any cheaper than actually going out to get your own fresh food if I’m honest.* (Participant 14)

A few rejected the idea that UPFs are necessarily cheaper.

> *I would say it’s cheaper to make something by yourself but it also depends the ingredients you’re gonna use, because if you want to cook something with only organic product it’s gonna be same or maybe bit more expensive but if you just use simple ingredients you can make a meal much cheaper than buying something or ordering online.* (Participant 4)

When the price difference between cooking from scratch and buying UPF was close, some participants suggested convenience may be the deciding factor.

> *A couple of days ago I bought a pizza and last week I made my own pizza, so I would say making your own pizza is cheaper, I think it’s just time, it’s the time that you just make an order, you go and pick it up in 10 minutes, case closed.* (Participant 3)

For others, the cost premium of some types of convenience was sometimes considered unaffordable.

> *Recently I ordered one takeaway home; this is only as a treat once a month, maybe twice a month. I remember last year it was much cheaper, but these days with the cost-of-living crisis and everything, the price I paid was huge, and that put me off for ordering takeaways, probably forever.* (Participant 6)

Alongside trading off convenience against food costs, some participants also noted the trade-off between the short-term benefits of UPFs and the long-term health harms.

> *In the short [term] you probably save time and save some money, but in the long-term you’re actually exposing yourself to illnesses.* (Participant 2)

#### Food culture and cooking skills

Some participants described how cultural values, food knowledge and skills, and family traditions helped them avoid UPFs. Growing up in households where home-cooked meals were the norm helped some participants continue those habits into adulthood. This was often supported by perceptions of strong cooking skills, time availability, or confidence in interpreting health information. However, others described a gradual erosion of these protective factors across generations. Those who lacked this foundation expressed more resignation toward UPFs.

> *I think in terms of my parents, [their way of eating] is now more habit, they have done it since childhood, why change when they are in their sixties? And they are still alive… this is the type of thing my mum would say is, ‘well it hasn’t killed me up until now, so why would it now? It doesn’t matter what they say’.* (Participant 25)

Some participants reflected on how cultural shifts had normalised UPF consumption, particularly in the UK, where pre-packaged and takeaway foods were seen as standard options. Some participants noted that moving to the UK had significantly altered their diet, shifting them toward greater UPF consumption.

> *I think in Greece we have our own fast food, which might seem a bit healthier to an extent but I don’t know how much it is… the fast food is much more spread here in terms of burgers and pizzas and I think that probably plays another role… I didn’t start eating burgers before I came here 5 years ago, it wasn’t that popular in Greece.* (Participant 3)

Others described generational differences, with younger generations increasingly reliant on ready-made foods due to lack of cooking skills and exposure to convenience culture.

> *When I was younger my mum used to cook and we all used to eat so that carried on to me, I cook, my children eat, and then I can’t understand the ones that don’t cook and just stick pizzas in the oven… I know there’s parents out there whose lifestyle is completely different where they would bring their children home from school, stick them in front of the TV and then give them their processed food, but that’s not because mum was working, that’s just because that’s her way of life.* (Participant 12)

### Theme 5: Proposed solutions for reducing UPF consumption

Participants expressed a range of views on potential interventions to reduce UPF consumption, including government regulation, education, and voluntary industry action. While some supported policies such as taxation or marketing restrictions, others expressed concerns about overreach.

#### Marketing restrictions

Participants expressed support for strong government policies to reduce UPF consumption, particularly among those who viewed diet as a major contributor to population ill health.

These participants often argued that responsibility for change should not fall solely on individuals, and that governments should intervene directly. Some called for tighter restrictions on the content and marketing of UPFs.

> *I think if governments all over reined in what is and isn’t allowed in food, it would also see this mass health crisis that we’re going through reined in as well… health services are inundated with cardiac problems and obesity… and in the middle of it all is processed foods… Look at child obesity figures. Look at the increase in cancer figures. Look at the massive increase in liver diseases… You look into that and the source is all pointing towards diet… I would just outright block them, absolutely put heavy restrictions on what can go into food, how food can be marketed, who it’s marketed at … a lot of it’s aimed at kids, teenagers, busy parents, parents who claim they can’t cook, just whoever is looking for convenience… I don’t think we should be encouraged by advertisements to poison ourselves.* (Participant 17)

However, participants also expressed despondency over whether it would be possible or effective to restrict UPF marketing.

> *We are a capitalist environment, so I don’t know how we can restrict marketing really because whatever we like to buy we will find it in the supermarket, so it’s like whatever we have available is indication of what we want to buy.* (Participant 4)

#### Taxation and financial incentives

Participants referenced the UK’s Soft Drinks Industry Levy (or ‘sugar tax’), suggesting a similar approach could reduce UPF consumption. Some supported financial incentives and subsidies for minimally processed foods instead. Others expressed concern that taxes could fuel resentment, especially if seen as limiting choice or unfairly impacting those with fewer food options.

> *[the] sugar tax I think does help encourage people to avoid that much sugar, and I think likewise they can do that on ultra-processed foods… I guess it would definitely allow people to prefer the more healthy option because it’s cheaper, but I guess like what you said, people who can’t afford it, it might restrict what they can indulge in, which sometimes some people might think of that as like being unfair.* (Participant 13)

#### Public messaging, education, and the need for clarity

Some participants questioned whether dividing food into different levels of processing was useful, suggesting that public messaging might be more effective if focused on more intuitive concepts rather than technical classifications.

> *Whether calling them UPF foods is helpful or not, I would more or less immediately say no, it’s very unhelpful, because people can probably understand the basic generalisation of processed foods, but then when you’re taking it that one step further and adding all those complications into the definition, I think Joe Public would probably shut off at that point because they don’t need to know. That makes too much information.* (Participant 15)

While many participants supported education as a tool for reducing UPF consumption, particularly among children, comments reflected a desire for simplified, practical guidance.

> *First of all, if you target the young generation first as like what you said education. So like in school, actually teach students about what ultra-processed foods mean and how to tell the difference… teaching kids early in terms of what a healthy diet is.* (Participant 13)

## DISCUSSION

### Summary of main findings

This is one of the first studies to provide an in-depth exploration of public understandings of the concept of UPFs, influences on these perceptions and how these affect food decisions. We conducted 30 interviews with UK adults with household responsibility for food. Interviews revealed a spectrum of knowledge about UPFs. Food processing often made more sense to participants as a continuum rather than ordinal categories. There was particular confusion regarding the boundary between processed and ultra-processed food. Simple shortcuts were often used to identify UPFs such as ‘single ingredient foods’.

Participants described overwhelming information including from traditional media, social media, and the experiences of family and friends. It was not possible to draw causal connections between individual perceptions and information sources. Rather, multiple information sources interacted in a cacophonous media environment which included both traditional and online-only sources. Participants recognised the potential for misinformation, but often the same sources were considered to provide both mis- and trusted information. Social media was particularly dominant, potentially because it offered a constant stream of commentary and recommendations, unlike the more episodic nature of books or television programmes.

Personal and anecdotal experience was particularly powerful and many described trial and error processes for exploring dietary effects on their wellbeing. This was the case at both ends of the spectrum: people who had eaten UPFs their whole lives were reassured they had survived up until this point, whilst those trying to gain control of health problems experimented with changes. Neither of these groups thought of risk in terms of epidemiological averages but viewed it through the lens of their own and others’ experience. When personal experience conflicted with professional advice, the latter was often seen as outdated, and participants were aware of scientific uncertainty around the health effects of UPFs. While industry-sponsored information was met with scepticism, views on social media influencers were more mixed, and their personal testimonies were sometimes seen as relatable or helpful.

Participants’ perceptions of UPFs sometimes influenced their purchasing and consumption, particularly among those managing diet-related conditions. However, many participants described structural barriers as primary reasons for purchasing decisions. Cost, convenience, taste, and family practices were key determinants of food choices. Finally, a diversity of approaches was offered to help reduce UPF consumption, from education to fiscal policies.

These findings suggest that the public need help navigating the complex food information landscape and not just with filling specific knowledge gaps around UPFs or other topics. Calls for education may reflect a desire for trusted, simplified tools to help people make better food choices in an environment saturated with both marketing and scientific complexity.

### Strengths and limitations

We took several steps to enhance the rigour and trustworthiness of the research. PIE contributors were consulted during the interpretation of findings, to ensure that themes and illustrative quotes were considered through diverse and grounded perspectives. Team-based analysis included a data clinic with multiple researchers and double-reading of transcripts to support reflexivity and consistency in interpretation. While the sample was not intended to be statistically representative, we achieved diversity in age, gender, education, and geographic location. Most participants were women and more highly educated, which likely reflects the inclusion criterion of being primarily responsible for food decisions, the recruitment strategy via social media, and differential willingness to engage in food-related research. It is also possible that individuals with personal interest in diet and nutrition, or diet-related concerns, were more likely to participate. Some groups, particularly those with less formal education and Black people, were underrepresented. These limitations may affect the transferability of findings, particularly to individuals or communities experiencing food insecurity, where structural drivers of UPF consumption may be even stronger (42). Nonetheless, our aim was to explore a broad range of perceptions, and the findings are likely transferable to other high-income countries with similar food environments and levels of UPF availability.

### Interpretation of findings and relationship to prior literature

While previous research has primarily used survey data or focus groups to explore public perceptions of UPF (17,43), this in-depth qualitative study contributes novel insights into how individuals engage with the wider information environment around UPFs, the challenges they face in interpreting the concept, and the information sources that shape their perceptions. By capturing the varied ways that people navigate the complex information environment and relate it to their dietary choices, this study reveals key barriers and opportunities to enhance public understanding of UPFs.

Our finding that many participants struggled to define or apply the concept of UPFs aligns with previous studies showing inconsistent or inaccurate understanding of the term. For example, a recent survey in Brazil found that although over 80% of respondents had heard the term UPF, the majority defined it broadly as “foods that have gone through many processes” and often misclassified items such as yoghurt, cereal bars, and frozen vegetables (34). We also found evidence of confusion on the borders between processed and ultra-processed. In contrast, research among trained nutrition professionals has shown good inter-rater reliability when structured training on Nova classification is provided (31), suggesting that while the framework is internally coherent, it may be challenging to apply without training. Some of this confusion may stem from how UPFs are framed in the media, with evidence from Australia showing that industry communications often conflate foods from Nova groups 3 (processed foods) and 4 (UPFs) (26).

Our findings regarding why people reported consuming UPFs align with previous literature, with key determinants including affordability, convenience, availability, cultural norms, taste preferences, and marketing influences (44). However, convenience was consistently described as one of the key reasons for eating UPFs. Survey data on public perceptions of healthy eating in the USA similarly indicates that convenience is a key consideration in food choices (45).

In some cases, personal experiences of chronic conditions shaped how participants evaluated dietary advice. Those who felt mainstream medical recommendations had failed to address their health concerns were more likely to seek information elsewhere and express scepticism toward traditional experts. Chronic conditions can be difficult to treat, causing people to seek alternative sources of information (46–48). Other research has similarly found that people are more likely to exhibit a ’backlash’ to conflicting nutrition information from traditional experts than to conflicting information from other sources (49). This may reflect not only a lack of trust, but also frustration with the complex, inconsistent, and often overwhelming nature of navigating the nutrition information landscape. Participants in our study frequently expressed a desire for clearer and more actionable information, suggesting that improving the simplicity, quality, and consistency of communication about UPFs could be more helpful than disseminating scientific information or technical category boundaries. More general guidance on how to navigate cacophonous information environments, a phenomenon not limited to the diet and food space, may also be helpful.

This study also adds to the literature on the acceptability of dietary public health policies, revealing varied public attitudes toward potential UPF-related interventions. While some participants supported regulatory measures like taxation or marketing restrictions, others expressed scepticism about the need for these or concerns about personal autonomy and regressive impacts. These findings align with evidence that policy acceptability is associated with public understanding of the rationale behind interventions and their perceived fairness (35).

Finally, our findings suggest that public awareness of the commercial determinants of health, particularly industry influence and marketing tactics, may be increasing. While traditional media has historically shaped food narratives, the rise of new online-only media platforms, such as social media, has not only amplified conflicting messages but may also be contributing to greater public scrutiny of food industry practices. In this context, participants described an information landscape where the lines between credible guidance, commercial marketing, and personal anecdote are increasingly difficult to identify. This aligns with emerging literature on the commercial determinants of health, which highlights how marketing strategies influence dietary norms, consumption patterns and health outcomes (50).

### Implications for research, policy and practice

Given the nature of the participants we recruited and our conclusion that results may not be transferrable to those experiencing food insecurity, future research should make efforts to explore these topics in groups underrepresented here. Survey research could be used to quantify the prevalence of different perceptions described here overall and across different demographic groups.

Whilst education on the nature of UPF could improve public understanding, our findings indicate that support with navigating complex information environments and identifying mis- and disinformation may be more helpful to individuals trying to make sense of what and how they should eat. Policy interventions to restrict marketing, availability and affordability of UPFs, as well as increase affordability of non-UPFs, would be welcomed by some.

Convincing and consistent messaging of the harms of UPFs may help increase public support for these policies. Future research could explore how to support individuals in navigating these information environments, including through improved media literacy and the ability to critically evaluate information (22,51), and through clearer communication about the health risks of UPFs and the marketing strategies used to promote them (23,25,49).

## CONCLUSION

This study reveals how UK adults with household responsibility for food activities makes sense of UPFs in a complex information environment. Although we identified a range of public understanding of UPFs, several barriers to reducing consumption were consistently identified, including confusion over definitions, the convenience and affordability of UPFs, time poverty, and the ubiquity of ready-made foods. A strong food culture that included cooking skills and traditional eating patterns helped people avoid UPFs. Participants also expressed a desire for simpler, more practical information and structural support to make healthier choices. These findings underscore the need for policy approaches that combine improved public communication with upstream interventions that affect the food environment to make choosing less processed foods easier and more appealing.

## Supporting information

Supplementary File 1 COREQ checklist

Supplementary File 2 Participant Info Sheet

Supplementary File 3 Interview Topic Guide

## Data Availability

The datasets generated and analysed during the current study are not publicly available as it is difficult to fully anonymise qualitative transcripts.

## Ethics approval and consent to participate

The study received ethics approval from the Humanities and Social Sciences Research Ethics Committee at the University of Cambridge: reference 24.365. All participants provided written informed consent before data collection began.

## Consent for publication

Not applicable.

## Authors’ contributions

ME, JCP, EPV, MW and JA were involved in conceptualisation and design. ME was involved in data acquisition. ME, JCP, JA, ME, PS and DS were involved in data interpretation. ME and JA drafted the work. All authors contributed to substantive revisions of the work, have approved the submitted version and agree to be personally accountable for the work.

## Acknowledgements

Not applicable.

## LIST OF ABBREVIATIONS

PIE: public involvement and engagement
UK: United Kingdom
UPF: ultra-processed food
USA: United States of America

