## Supplementary File 2 Participant Info Sheet for "Understandings of ultra-processed foods: a qualitative interview study of UK adults with responsibility for household food activities"

### Participant Information Sheet

### Perceptions of the concept of "ultra-processed foods" in UK adults

#### Summary

Ultra-processed foods are being covered by the news more frequently, due to the recent release of books, TV programmes and podcasts on the topic.

We want to understand your experiences and understanding of the term ultra-processed foods, or UPFs. We are particularly interested in whether you have heard of the term from any form of media, how you understand the term, and whether your understanding of the term is related to the foods that you buy and eat.

Please take the time to read the following information carefully. Discuss it with family and friends if you wish and decide whether you wish to take part. Thank you for your support with our research so far.

#### Contents

1 Why we are doing this study
2 Why am I being asked to take part?
3 What will happen to me if I take part?
4 Possible benefits and disadvantages of taking part
5 More information about taking part
6 Contact for further information

#### How to contact us

If you have any questions about this study, then please talk to:

**Dr Michael Essman**
MRC Epidemiology Unit
University of Cambridge School of Clinical Medicine
Box 285 Institute of Metabolic Science
Cambridge Biomedical Campus
Cambridge CB2 0QQ


#### 1 Why we are doing this study?

##### What are we studying?

In this research we are aiming to interview adults with responsibility for food shopping and preparation in their households to explore their understanding of the concept of UPF and its relationship to health.

We will ask whether you have heard of the term ultra-processed foods—from the media or elsewhere—and whether what you have heard about UPFs affect how you decide what foods to buy.

We will share our results with people working in Government, other researchers, and the public. Our results could help Government understand public opinion about UPFs and whether they should enact any policies aimed at UPFs.

#### 2 Why am I being asked to take part?

We are inviting you to take part because you have said that you are usually the main person in your household in charge of activities related to food, for example shopping and preparing food.

#### 3 What will happen to me if I take part?

We will conduct a **one-to-one interview with you via Microsoft Teams** to better understand your experiences with and understanding of ultra-processed foods. You will only be asked to complete one interview and it will last a maximum of 60 minutes.

Before taking part in an interview, you will have the opportunity to speak with a researcher and ask questions about the study. If you are willing to take part, you will be asked to provide consent to take part using an online form (e-consent).

The interviews will be audio-recorded to help us retain key insights shared. Quotations that contain no identifiable information may be used in research publications resulting from the interviews. No one will have access to the recording or any information about you except members of the research team at the University of Cambridge.

#### 4 Possible benefits and disadvantages of taking part

##### What are the possible benefits of taking part?

You will help us understand how members of the public understand the concept of UPFs, and this could help with the development of future healthier eating policies, both through the contribution of these interviews to academic work and to interested policymakers in Government.

##### What are the possible disadvantages and risks of taking part?

Taking part in this study will involve sacrificing some of your time.

#### 5 More information about taking part

#### Do I have to take part?

No, it is up to you to decide whether to take part. You are free to withdraw at any time, without giving a reason.

#### Will I receive any payment for taking part?

We will offer a £20 Amazon gift voucher for one interview to you as a gesture of thanks and recompense for your time.

#### What if there is a problem?

If you have a concern about any aspect of this study, you should ask to speak to the research team who will do their best to answer your questions. If you remain unhappy and wish to complain formally, the University of Cambridge complaints process is available to you through the University of Cambridge Clinical School Secretary: telephone: 01223 333543 or

#### What will happen to information about me collected during the study?

The interviews will be video-recorded and transcribed using Microsoft Teams. All names will be removed from the transcript, and it will only be identified by an anonymised code. No one will have access to the recording except members of the research team at the University of Cambridge.

Information we collect during the research will be kept strictly confidential. Any information about you will have your name and contact details removed so that you cannot be recognised from it and it will not be used or made available for any purpose other than for research. We may used anonymised quotations from the interviews in reports and publications on the study.

With your permission, information will be stored anonymously at the MRC Epidemiology Unit on a secure research drive. Codes connecting your individual identity to the stored data records will be kept separately. The database containing personal information is on a secured network drive on computers in the MRC Epidemiology Unit, University of Cambridge.

Occasionally our studies may be monitored by our Sponsors. This is to ensure our research is conducted soundly. This procedure is routine and carried out by fully qualified personnel and data confidentiality will be always adhered to. At the end of the study the confidential records will be kept for a maximum of 20 years and then destroyed.

The University of Cambridge is the sponsor for this study based in the United Kingdom. We will be using information from you to undertake this study and will act as the data controller for this. Cambridge University will keep identifiable information about you for a minimum of 20 years after the study has finished.

Your rights to access, change or move your information are limited, as we need to manage your information in specific ways for the research to be reliable and accurate. If you withdraw from the study, we will keep the information about you that we have already obtained. To safeguard your rights, we will use the minimum personally identifiable information possible.

You can find out more about how we use your information at https://www.information-compliance.admin.cam.ac.uk/data-protection/applicant-data

#### What will happen to the results of the study?

When the study is completed, the results will be published in an academic journal without a paywall so that anyone can see the results. We may also present the results orally at scientific meetings and to interested stakeholders, including representatives from the UK Government's Department of Health and Social Care. If published or presented, your identity, affiliation and personal details will be kept confidential. No information that could identify you, like your name, will be published in any report about this study.

#### Who is organising and funding the study?

This study is organised by the MRC Epidemiology Unit, part of the University of Cambridge. The study is funded by the National Institute for Health and Care Research School for Public Health Research. We are working with Imperial College London. This study is one part of a larger research collaboration with Imperial College London that is trying to understand how UPFs are presented in the media and understood by the public.

#### Who has reviewed the study?

This trial has been reviewed by an independent group of people, called a Research Ethics Committee, to protect your safety, rights, wellbeing, and dignity. The study has been granted approval by the University of Cambridge Humanities and Social Sciences Research Ethics Committee. You can contact the Committee by quoting the ethics reference 24.365 by

### 6 Contact for further information

If you have any questions regarding the study or how you might be involved further contact information can be found below.

#### Study Lead

**Dr Michael Essman**
Research Associate
MRC Epidemiology Unit
University of Cambridge


#### Principal Investigator

**Prof Martin White**
Programme Leader and MRC Investigator
MRC Epidemiology Unit
University of Cambridge


#### Thank you for taking the time to consider taking part in this study.
