## Supplementary File 3 Interview Topic Guide for "Understandings of ultra-processed foods: a qualitative interview study of UK adults with responsibility for household food activities"

**Data collection for: Perceptions of the concept of “ultra-processed foods” in UK adults**

**Interview topic guide**

*Note: use separate topic guide for each interview.*

**Interview #:______________**

Interview conducted via Microsoft Teams

**Pre-Interview: Participant information, screening question, and signing consent form**

Before we begin the interview, I want to make sure you have had the opportunity to review the participant information sheet and ask questions.

As a reminder, for this study we are interested in interviewing people who are in charge of the buying and preparing of food in their household. Are you usually the main person in your household in charge of activities related to food, for example buying and cooking? **(screening Q)**

Are you happy that you understand the research project and what it will entail? [*Let them answer]*

Once you are ready, we can sign the online consent form and proceed to the interview. [*Cannot proceed without completing written E-consent]*

**Interview: Introduction**

Thanks for agreeing to take part in an interview for our study today. This interview is expected to take up to a maximum of 60 minutes. Is that OK with you, and do you need to finish earlier for any reason? Are you in a comfortable space to conduct the interview and not be interrupted? [*Can reassure them interruptions are OK, just want to make sure they’re ready and comfortable]*

My name is Mike Essman, I work with the MRC Epidemiology Unit at Cambridge University. Before we start on the interview, I’m going to briefly remind you of the topic we are going to discuss today. We want to understand your experiences and understanding of the term ultra-processed foods, or UPFs. We are particularly interested in whether you have heard of the term from any form of media, how you understand the term, and whether your understanding of the term is related to the foods that you buy and eat.

As a reminder, all questions are optional, and there are no right or wrong answers. We are only interested in your experiences and thoughts, and we can skip any questions that you do not feel comfortable answering.

Do you have any questions before we start? [*Let them answer]*

I will now start the recording. [START RECORDING ON TEAMS]

*Discussion begins (below), make sure to give people time to think and don’t move too quickly. Use the probes to make sure that all issues are addressed, but move on when you feel you are starting to hear repetitive information and allow for flexibility in the direction that the participant takes the conversation.*

| **Check** | **#** | **Question** | **Probe** |
| --- | --- | --- | --- |
| **Introduction / Screening Question – overall role of household food purchases** | | | |
| **Could you confirm your age, gender, ethnicity, highest education attained, and the county where you live? (hoping to interview a range of ages and gender)** | | | |
| ☐ | 1 | What is your role in buying and preparing food in your household? | -What are some of the main things you think about when buying or preparing food? |
| ☐ | 2 | Have you ever heard of the term ultra-processed foods, or UPFs, and can you remember where you heard it? (proceed with using either full name or acronym for UPFs depending on which participant prefers) | -How would you explain what UPFs are, and how would you recognise them?  -Have you heard about ultra-processed foods or UPFs from any other sources? Maybe from friends, colleagues, the news, podcasts, or other media? |
| ☐ | 3 | Next, we are going to take a look at a few different types of foods to discuss whether they seem ultra-processed or not, and why you think so. | *Examine photo slides of UPFs and less processed foods to make sure interviewee understands what they are [only possible on video conference, not phone call. If they do not have Teams, may just need to describe them to get a sense of what UPFs are.* |
| **Perceptions of UPFs, how much do they purchase, and why** | | | |
| We are interested in your opinions about and experiences with UPFs: how you understand the term and what you think about them. | | | |
| ☐ | 4 | What comes to mind when you think about UPFs? | -What does ultra-processed food mean, and how is that different from processed food?  -What are the most important considerations for you when buying food? (e.g. taste, time spent, healthiness, or high/low in certain nutrients)  -What characteristics do you think of when you look for healthy food? |
| ☐ | 5 | Making your best guess, about how much of the food that you purchase for the household during a typical week would be considered ultra-processed? | -If it’s easier, we can talk through typical breakfast, lunch, dinner, and snacks.  -If needed, prompt: very little, some but less than half, about half, more than half, nearly all |
| ☐ | 6 | Do you think there are any benefits of buying or eating UPFs? | -Do UPFs make your life easier or more enjoyable in any way? How? What about for others in your household? How? Why?  -Do UPFs help you with your food purchasing and/or cooking tasks? How? Why?  -Do you think UPFs are cheaper or more convenient than less processed foods? How? Why?  - Are they tastier? How? Why? |
| ☐ | 7 | Do you think there are any harms or negative aspects of buying or eating UPFs? | -Are there any downsides of buying or eating UPFs? How? Why?  -Does it matter if you eat them frequently or infrequently? How? Why?  - Do they make shopping or cooking harder? How? Why?  - Are they more expensive or less convenient? How? Why?  - Are they less tasty? How? Why?  -Is it more/less enjoyable if you consume more/less? |
| ☐ | 8 | If you wanted to purchase unprocessed or minimally processed foods, how would you recognise them? | -Are there certain characteristics about unprocessed or minimally processed foods that make them easy to identify? (e.g. related to packaging, labels)  -Is this easy or difficult to do and why? |
| ☐ | 9 | Would you say that UPFs are healthier, less healthy, or neither more nor less healthy than foods with less processing? (*this may have come up in Qs 6&7 but keeping to make sure*) | -Are there certain aspects of processed foods that are good for you?  -Are there certain aspects of processed foods that are bad for you? -Do you think eating UPF are compatible with eating a healthy diet? |
| **Sources of information regarding UPFs and associations with health** | | | |
| Thank you for your responses so far. We are interested in some of the media sources from which you may have heard about UPFs. Could you tell me a bit more about where you have heard about UPFs and what you can remember was said about them? | | | |
| ☐ | 10 | From what sources have you heard about UPFs? These could include television, online websites or news stories, friends, podcasts, magazines or other media. | -[*Probe for more sources*] Are there any sources you think you may have left out from before? Sources could include friends, colleagues, social media, the news, podcasts, or other media? |
| ☐ | 11 | Thinking about the sources of information we just discussed, do you trust those sources? Why or why not? | -Do you agree or disagree with these statements about UPFs?  -Did the source say anything about the health effects of eating UPFs?  -Which sources do you consider more credible and influential when deciding whether to take their messages about UPFs on board? [may need prompting about the different types of sources friends/family vs. scientists vs. general media and so on]  -Which of the sources you’ve mentioned do you trust the most and which do you trust the least?  -In your opinion have you noticed a significant shift in people’s use of the term UPFs recently? [probe how supermarkets, restaurants may be presenting food offerings; and also how friends/family are using UPFs and non-UPF in daily decisions about food]  -Have you sought our more information about UPFs on your own, and if so, from where? |
| ☐ | 12 | Has the information you have heard about UPFs led you to eat more or less of them, and if so, why? How have you changed how much UPF you buy/eat/provide for your household recently, and why? | -Probe for the different reasons why they might/might not want to change their eating of UPFs |
| ☐ | 13 | Would you support a Government policy that aimed to reduce how much UPFs people eat? Why?  What types of policies do you think could work to change how much UPFs people in England eat?  Thinking about your own household, which policies do you think could change the amount of UPFs your household consumes? Why? | -What would make these policies more/less likely to be effective?  -What would be the potential benefits or harms of such a policy for you or others? |
| **Concluding remarks** | | | |
| ☐ | 14 | That’s all the questions I have for you today, thank you again for your contribution. *Briefly review discussion, ask any questions I missed or didn’t get full answers to.*  Do you have any questions, or is there anything we haven’t covered which you think might be important in our research before I switch off the recorder? |  |
| ☐ | 15 | Also, do you have friends or colleagues that you would recommend we speak to? *Read inclusion criteria to them.* | ****Add names and contact details to longlist after interview**** |
| Thank you for your time. I’m going to switch off the recorder now. [SWITCH OFF AUDIO RECORDER] | | | |
| **Data collection diary** | | | |
| *Add anonymous comments only, to aid interpretation of transcripts – store in SRD* | | | |

**Post-interview checklist for interviewer:**

**☐** Audio recording checked that the voices are clear, and transcription has been generated by Teams.

**☐** Edited/corrected Teams transcript as necessary and anonymised participant identity.

**☐** Audio recording uploaded to SRD.

**☐** Once safely transferred to the SRD, all files deleted from TEAMS.

**☐** *If applicable* add contact(s) offered for snowball sampling to the interview contact list.

****INTERVIEWER MUST UPLOAD THIS DOCUMENT TO SRD UPON COMPLETION****
